# Seroprevalence and risk factors for SARS-CoV-2 infection in middle-sized cities of Burkina Faso: a descriptive cross-sectional study

**DOI:** 10.1101/2024.03.13.24304208

**Authors:** Adama Sana, Elodie Djemaï, Philippe De Vreyer, Thomas Thivillon, Hermann Badolo, Abdramane Berthé, Dramane Kania

**Author notes:** Corresponding author: Elodie Djemaï. These authors contributed equally to this work (Adama Sana, Elodie Djemaï). These authors also contributed equally to this work (Hermann Badolo, Abdramane Berthé, Dramane Kania).

## Abstract

**Background:** Since March 2020, COVID-19 has evolved from a localized outbreak to a global pandemic. We assessed the seroprevalence of COVID-19 in three towns in the Centre Sud region of Burkina Faso.

**Methods:** A population-based cross-sectional survey was conducted in three medium-sized towns in Burkina Faso’s Centre Sud region, from June to July 2021. Subjects aged 16 or over at the time of the survey were considered for this seroprevalence study. The Biosynex COVID-19 BSS rapid test was used to detect immunoglobulin G (IgG) and immunoglobulin M (IgM) against SARS-CoV-2. A standardized questionnaire was also administered to collect additional information.

**Results:** A total of 2449 eligible participants (age ≥ 16 years) were identified. Serological tests for COVID-19 were performed in 2155 individuals. Finally, 2143 valid tests were retained and analyzed. Out of the entire sample, 246 positive tests were observed, corresponding to a prevalence of 11.48%. Prevalence was 9.35% (58 cases) in Kombissiri, 12.86% (80 cases) in Manga and 11.99% (108 cases) in Pô. By gender, 13.37% of women (164 cases) tested positive, and 8.95% of men (82 cases). Women accounted for 66.67% of all positive test subjects. The results show a significantly higher seroprevalence in women (P = 0.007), people over 55 years old (P = 0.004), overweight or obese people (P =0.026) and those with drinking water sources at home (0.013).

**Conclusions:** The results of this study show that the COVID-19 virus also circulates in the population of medium-sized towns in Burkina Faso, far more than is officially reported in the country. The study also highlighted the greater vulnerability of women to the epidemic, and the challenge of access to water in the face of diseases such as COVID-19. The preventive measures put in place to fight the pandemic must take these different factors into account.

## Introduction

Coronavirus disease 2019 (COVID-19) is an infectious disease whose initial cases were first reported in Wuhan, Hubei Province, China, in December 2019 [1]. Rapidly evolving from a localized outbreak, it transformed into a global pandemic [2].

Burkina Faso, like many other countries worldwide, was also affected. The country recorded its first confirmed case of SARS-CoV-2 infection, the virus responsible for COVID-19, on March 9, 2020. As of July 3, 2022, there were officially 21,134 confirmed COVID-19 cases in the country, with 387 deaths [3]. The capital-city, Ouagadougou, situated in the Center region, and Bobo-Dioulasso, the country’s second-largest city located in the Hauts-Bassins region, were the main affected areas, and benefited the most from response measures.

Data from the countries earliest affected by the pandemic indicate that approximately 40% of infected individuals will exhibit a mild form of the disease, often asymptomatic [4]. Despite the absence or presence of mild signs and symptoms, these individuals are contagious and thus capable of transmitting the disease to others [4].

Fragility of healthcare systems, limited access to hygiene and sanitation facilities, and lack of early treatment options during the initial stages of the pandemic raised concerns about a potentially rapid increase in infections in sub-Saharan Africa. Governments had few strategies available other than slowing down the virus transmission as much as possible. This study aims to assess the epidemic situation in medium-sized cities in the Center-South region of Burkina Faso and to identify the risk factors for COVID-19 infection.

## Materials and Methods

### Study Design

This study was a descriptive cross-sectional investigation based on survey data collected from June 7 to July 3, 2021.

### Study Site

The study was conducted in three cities in the Center-South region of Burkina Faso: Kombissiri, Manga, and Pô.

### Study Population and Sampling

The sample of households comes from a list of households that had been surveyed between November 2019 and March 2020 for a study on domestic cooking fuel choices and exposure to air pollution. The households were initially randomly selected using a spatial sampling strategy, with GPS points drawn in the three study locations. Data collectors followed a predetermined random walk to select one household per GPS point.

The analysis relies on 813 households. 739 were already surveyed before the COVID-19 and 74 are replacement households.

### Inclusion and Exclusion Criteria

Eligible households were those without access to gas or electricity for cooking. This is a reasonable restriction as most households in Burkina Faso rely on solid fuels for cooking. Using the living standard measurement surveys collected in 2014 [5], it turns out that 83% of the households mostly rely on wood as primary cooking fuel at the national level, and only 13.4% have access to a cleaner cooking solution such as LPG or electricity. In addition, households declaring dolo brewing as one of their income-generating activities or reporting no cooking at home were excluded from the study.

### Consent

The project obtained approval from the Burkina Faso Health Research Ethics Committee on July 1, 2020, and from the International Review Board of the Paris School of Economics on March 16, 2021.

An information form outlining the terms and conditions of the research was read and explained to the members of the sampled households. Only after ensuring that the information contained in the information form is fully understood, is the participant’s consent requested by the data collector. To take part in the study, free, informed consent signed by the head of household or his/her representative was required. This consent was in both electronic (on tablet) and written (on paper) format. A signed and dated copy was given to the household before the start of the interviews. In addition, free and informed verbal consent was requested from each respondent. For minors, the consent of a parent or legal guardian was mandatory.

All participation was voluntary, and each participant was notified of his or her right to end participation at any time without having to provide a reason or suffer prejudice of any kind.

### Data Collection

As part of this study we administered questionnaires and did capillary blood sampling for rapid COVID-19 tests. The data collection started on June 7, 2021 and ended on July 2, 2021. Household members were interviewed using a standardized electronic questionnaire. Data collection covered sociodemographic characteristics, self-reported health, weight, height, blood pressure, and COVID-19 serological testing. Other variables were also studied and will be the subject of future publications.

For COVID-19 serological testing, a capillary blood sample was taken from study participants. The serological tests were only conducted among members of sampled households aged 16 and more. The rapid test used was the Biosynex COVID-19 BSS, an immunochromatographic test that detects anti-SARS-CoV-2 IgG and IgM antibodies. The theoretical performance provided by Biosynex was at least 96% sensitivity and 100% specificity.

### Data management and analysis

The SurveyCTO software was used for questionnaire programming and data recording on tablets, including serological test results. Data cleaning and analysis were performed using STATA (version 16.0). The analysis was primarily descriptive, with an analytical component considering sociodemographic variables that could potentially influence COVID-19 serological status.

## Results

### Sociodemographic characteristics of the study population

A total of 823 households were covered in the survey, encompassing 3,834 individuals, with 1,161 in Kombissiri, 937 in Manga, and 1,736 in Pô. Approximately 61% of households had more than five members. Serological testing for COVID-19 was conducted on 2,155 individuals out of the eligible 2,449 (aged 16 years and above). Among them, 12 tests were considered invalid due to the absence of the control line (C) on the cassette. Eventually, 2,143 valid and analyzed tests were retained, comprising 1,227 women and 916 men. Among the women, 38 declared being pregnant at the time of testing.

The analysis sample is made of 813 households in which at least one member was tested for COVID-19 during our survey and has a valid test. On average, there are 2.6 people tested by households (median 2, min 1, max 12). The mean age of those tested was 38.56 years (SD 17.46; min 16, max 111), with 38.76 years (SD 17.25; min 16, max 111) for women and 38.28 years (SD 17.74; min 16, max 92) for men. Approximately 56.40% of participants had never attended school. 41.06% lived in households with 1 to 5 members, and 6.71% had access to piped water (either inside their dwelling or from a neighbor). 66.04% had a normal body mass index (BMI), 15.83% were overweight, and 6.03% were obese (BMI ≥ 30). The descriptive statistics of the main variables are shown in Table 1.

**Table 1.** Descriptive statistics of the main sociodemographic variables.

| Characteristic | Frequency | Proportion (%) |
| --- | --- | --- |
| **Town (2143)** |  |  |
| Kombissiri | 620 | 28.93 |
| Manga | 622 | 29.03 |
| Pô | 901 | 42.04 |
| **Sex** |  |  |
| Female | 1227 | 57.26 |
| Male | 916 | 42.74 |
| **Age (in years)** |  |  |
| 16-24 | 599 | 27.95 |
| 25-54 | 1123 | 52.40 |
| ≥55 | 421 | 19.65 |
| **Body Mass Index (BMI)** |  |  |
| Underweight | 259 | 12.10 |
| Normal | 1414 | 66.04 |
| Overweight | 339 | 15.83 |
| Obese | 129 | 6.03 |
| **Education attainment (2108)** |  |  |
| No formal education | 1189 | 56.40 |
| Any formal education | 919 | 43.60 |
| Primary | 361 | 17.13 |
| Lower Secondary | 336 | 15.94 |
| Upper Secondary and above | 222 | 10.53 |
| **Household size (members)** |  |  |
| 1-5 | 880 | 41.06 |
| >5 | 1263 | 58.94 |
| **Access to water<sup>a</sup> (2131)** |  |  |
| Yes | 143 | 6.71 |
| No | 1988 | 93.29 |

Household members aged 16 or older whose RDT is valid were considered.
<sup>a</sup>Access to water is an indicator taking value 1 if the household has access to clean water in his house, garden or from a neighbor.

### COVID-19 Seroprevalence

Across the entire sample, 246 positive tests were observed, resulting in a prevalence of 11.48%. Prevalence was 9.35% (58 cases) in Kombissiri, 12.86% (80 cases) in Manga, and 11.99% (108 cases) in Pô.

Among the 246 positive cases, 164 were women (prevalence rate of 13.37%) and 82 were men (8.95%). Consequently, women constituted 66.67% of all subjects testing positive. Among pregnant women, the seroprevalence was 26.32% (10 cases out of 38).

The mean age of those testing positive was 41.22 years (SD 18.70; min 16, max 92). Seroprevalence was 10.51% (181 cases) in individuals under 55 years and 15.44% in those aged 55 years and older (65 cases).

### SARS-CoV-2 antibody prevalence and sociodemographic indicators

#### Univariate Analysis

We compared seropositivity according to city of residence, household size, sex, age group, education level, and access to water (Table 2). Univariate analysis from logistic regression did not show a significant difference between the three cities (P= 0.050 and 0.107), household size (P= 0.779), or education level (P= 0.790, a variable indicating whether one was educated or not, observed in 2,108 respondents). However, a statistically significant difference was noted for sex (P= 0.002), high BMI (P= 0.005, a variable indicating whether BMI is equal to 25 or more), age groups (P= 0.005), and gestational status among the 1,227 women (P= 0.021). Additionally, there was a significant difference based on whether a person was a member of a household with water access (private tap or shared tap in the courtyard or with a neighbor) compared to those without water access, with a p-value of 0.006.

**Table 2.** Odds ratio from univariate logistic regression.

| Variables | % Positive | Univariate OR | 95% CI | P-value |
| --- | --- | --- | --- | --- |
| **City (n=2143)** |  |  |  |  |
| Kombissiri | 9.35 | *Reference* | *Reference* |  |
| Manga | 12.86 | 1.43 | 1.00 2.05 | 0.050 |
| Pô | 11.99 | 1.32 | 0.94 1.85 | 0.107 |
| **Household size (n=2143)** |  |  |  |  |
| 1-5 | 11.26 | *Reference* | *Reference* |  |
| >5 | 11.65 | 1.04 | 0.79 1.36 | 0.779 |
| **Education (n=2108)** |  |  |  |  |
| No formal education | 11.32 | 1.04 | 0.79 1.36 | 0.790 |
| Any formal education | 11.69 | *Reference* | *Reference* |  |
| **Sex (n=2143)** |  |  |  |  |
| Female | 13.37 | 1.57 | 1.19 2.08 | 0.002 |
| Male | 8.95 | *Reference* | *Reference* |  |
| **BMI group (n=2141)** |  |  |  |  |
| BMI >25 | 10.46 | *Reference* | *Reference* |  |
| BMI larger than 25 | 15.17 | 1.53 | 1.14 2.06 | 0.005 |
| **Age (n=2143)** |  |  |  |  |
| 16-54 | 10.51 | *Reference* | *Reference* |  |
| 55+ | 15.44 | 1.55 | 1.14 2.11 | 0.005 |
| **Water access (n=2131)** |  |  |  |  |
| Yes | 18.35 | 1.82 | 1.19 2.79 | 0.006 |
| No | 11.00 | *Reference* | *Reference* |  |
| **Currently pregnant (n=1227)** |  |  |  |  |
| Yes | 26.32 | 2.40 | 1.14 5.04 | 0.021 |
| No | 12.95 | *Reference* | *Reference* |  |

Household members aged 16 or older whose RDT is valid were considered.
BMI = body mass index; n = number of individuals; OR = odds ratio; CI = confidence interval

#### Multivariate Analysis

In the multivariate analysis, we included all variables that had a significant association with SARS-CoV-2 infection (those with a critical probability below 0.10), except for gestational status, which was not included to analyze the complete sample of men and women. Logistic regression analysis revealed that female gender, high BMI, age (being under 55 years old), and lack of a household tap (private or shared) were factors associated with seropositivity to SARS-CoV-2 in our study population (see Table 3).

**Table 3.**
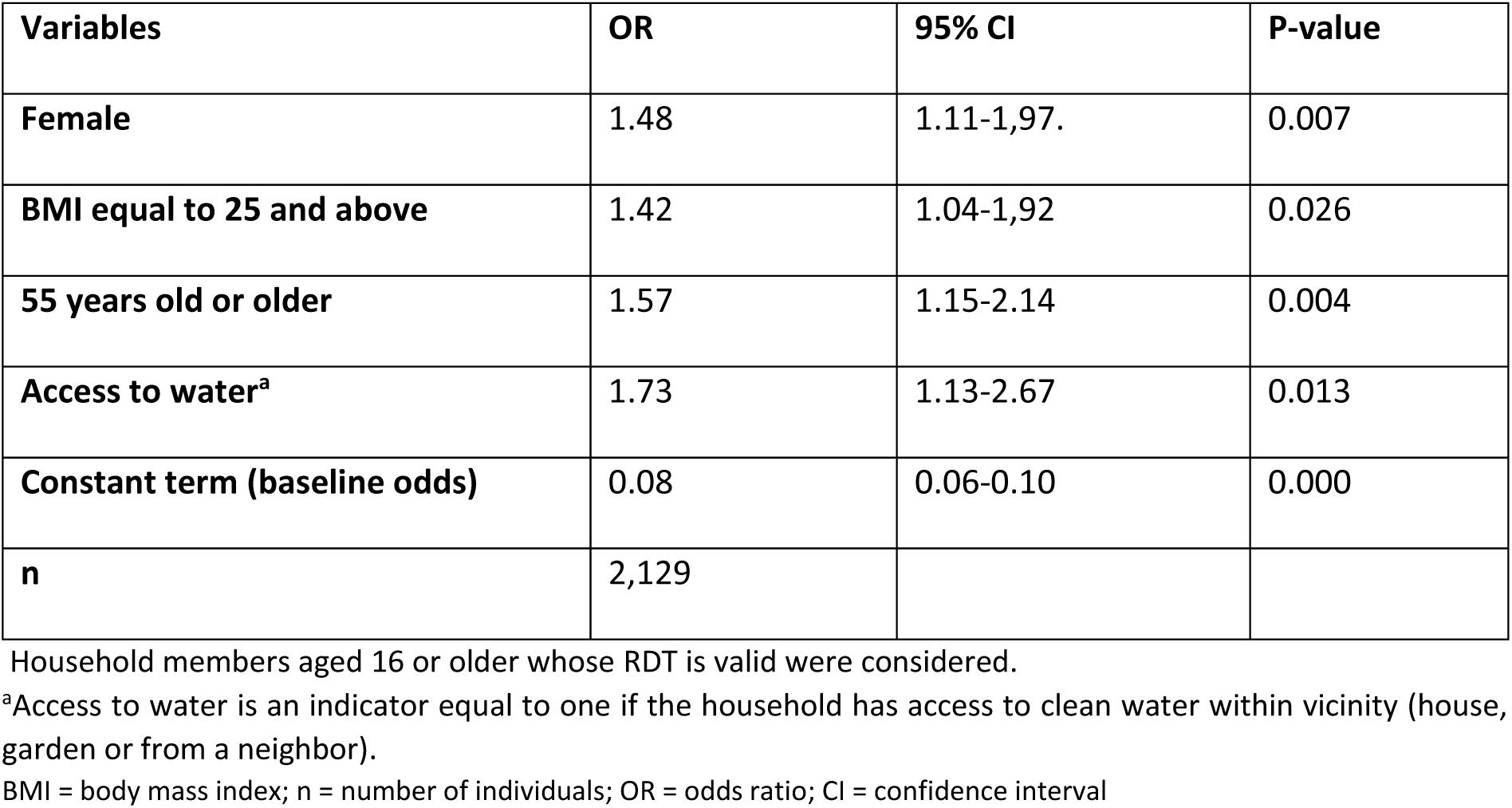
Odds ratio from multivariate logistic regression.

| Variables | OR | 95% CI | P-value |
| --- | --- | --- | --- |
| **Female** | 1.48 | 1.11-1.97. | 0.007 |
| **BMI equal to 25 and above** | 1.42 | 1.04-1.92 | 0.026 |
| **55 years old or older** | 1.57 | 1.15-2.14 | 0.004 |
| **Access to water<sup>a</sup>** | 1.73 | 1.13-2.67 | 0.013 |
| **Constant term (baseline odds)** | 0.08 | 0.06-0.10 | 0.000 |
| **n** | 2,129 |  |  |

Household members aged 16 or older whose RDT is valid were considered.
<sup>a</sup>Access to water is an indicator equal to one if the household has access to clean water within vicinity (house, garden or from a neighbor).
BMI = body mass index; n = number of individuals; OR = odds ratio; CI = confidence interval

## Discussion

This study demonstrates that the Central-South region of Burkina Faso was not spared by the COVID-19 pandemic. Despite the limited number of officially reported cases (25 confirmed cases from March 2020 to July 2021, and an additional 4 cases from August to December 2021)[3], our study reveals that the virus had circulated within the population. Sex, BMI, age, and the availability of a source of clean water within vicinity are factors significantly associated with seropositivity in our study population.

The average seroprevalence in the three cities under study (11.48%) is lower than that reported in Bamako, Mali, in September 2020 [6], where crude seroprevalence was 16.5% (225/1367, 95% CI = 14.5–18.5%), and in the Niger State (Nigeria), between June 26 and June 30, 2020 [7], with an average prevalence of 25.41%, including 21.43% in rural areas. In Africa, most prevalence studies found in our literature review were conducted on specific population groups, particularly those deemed to be at high risk of infection due to their activities or professions.

In Lomé, a study conducted from April 23 to May 8, 2020, on a group of individuals comprising healthcare workers, police officers, and airport personnel, reported a significantly lower prevalence compared to our study (1.6%; 95% CI: 0.9–2.6%) [8]. Similarly, in Kenya, the prevalence of anti-SARS-CoV-2 IgG antibodies among blood donors tested between April and June 2020 was 5.6% (95% CI: 4.8 to 6.5%) [9]. A study among blood donors in Quebec suggested that the low prevalence in the study population could be partly explained by self-exclusion bias [10].

Most of the seroprevalence estimations reported in the literature were conducted during the year 2020 (5-8), while our serological tests were conducted between June and July 2021. Burkina Faso experienced primarily two major waves of infection, the first between September and October 2020, and the second, which was larger, between late November 2020 and February 2021 (see Figs 1 and 2). A timid third wave was observed around June and September 2021. The COVID-19 vaccination campaign began on June 2, 2021, in the country.

**Fig 1.**
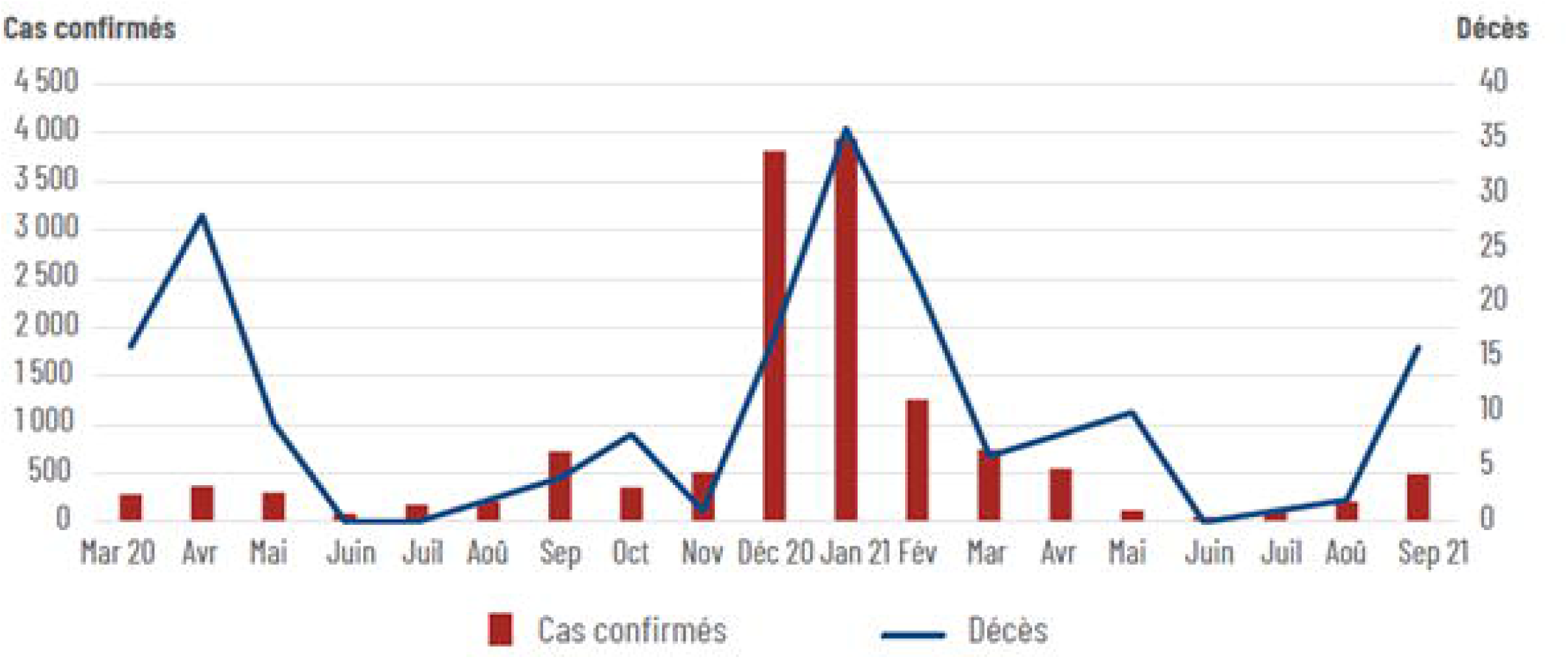
Number of confirmed cases and number of deaths due to COVID-19 infection between March 2020 and September 2021 in Burkina Faso. [11] Cas confirmés = confirmed cases Décès = deaths

**Fig 2.**
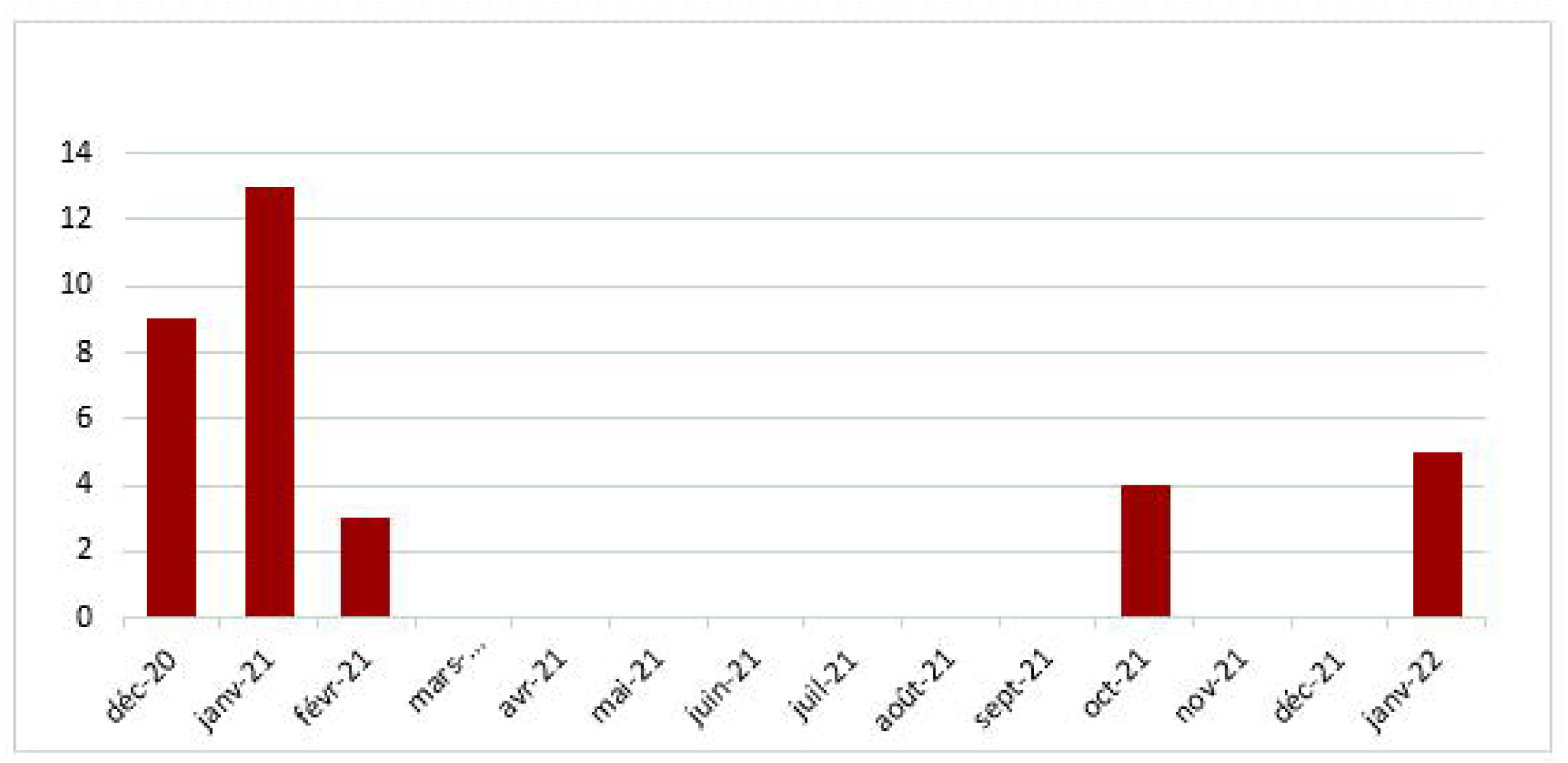
Number of confirmed cases of COVID-19 infection between March 2020 and January 2022 in the region of Centre Sud in Burkina Faso. *Calculations from Burkina Faso Open Data available on https://burkinafaso.opendataforafrica.org/jovpdge/burkina-faso-covid-19-rapport-de-situation [3]

The differences in prevalence between studies may be attributed, apart from differences in the composition of study populations (socio-demographic characteristics), to various factors such as the study period, the study setting (urban or rural), and the type of diagnostic test used (differences in diagnostic performance). Bobrovitz et al found that seroprevalence was low in general population compared to some specific populations and national studies had also lower seroprevalence than local studies [12].

In contrast to our study, Halatoko et al. reported a higher risk of SARS-CoV-2 seropositivity among individuals under 55 years of age compared to those aged 55 and above [8] and Bobrovitz found an higher seroprevalence among people ages 18–64 compared to 65 and over [12]. Similar findings were observed in the French SCOPE study, where the differences in the likelihood of infection were attributed to behaviors such as reduced outings and contacts among older adults [13]. Nevertheless, Nwosu et al. found that seropositivity rates progressively increased with age, ranging from 30.2% among individuals aged 15 to 29 years to 37.5% among subjects over 65 years [14].

These results could be explained by certain physiological predispositions that increase susceptibility to infections among older adults [14]. Moreover, a study conducted in France among participants aged 86 on average demonstrated that anti-Spike and neutralizing antibodies persisted for at least 9 months after SARS-CoV-2 infection in this age group [15]. Other researchers also revealed that individuals over 50 years of age or with a body mass index (BMI) greater than 25 had, one month after symptom onset, higher antibody levels compared to others, especially among men [16].

Our study indicates higher seroprevalence among females, consistent with findings from the SCOPE and EpiCov studies in France [17]. However, a systematic review of global seroprevalence of SARS-CoV-2 antibodies reported that there was no difference in seroprevalence between sex groups [12]. A study on gender inequalities in COVID-19 conducted in France identified a cross-effect of gender and socio-professional category. Women, due to their employment types, may face higher exposure to SARS-CoV-2 [18]. This prevalence difference between genders could also be attributed to women’s roles in certain communities, particularly in rural areas, where women care for the elderly, the sick, and engage in more frequent outings for visits and errands (e.g., water collection, market visits, income-generating activities) [18]. However, other research indicated that the duration of immunity against SARS-CoV-2 was longer in women than in men after infection [16].

Lastly, our study shows a relationship between having access to water within households or compounds and the likelihood of testing positive. We initially expected the opposite relationship, assuming that water access would lead to more frequent handwashing among household members. However, access to water was negatively correlated with poverty risk (p= 0.000) in our data, suggesting that greater access to water is associated with higher socioeconomic status. The observed positive relationship between access to water and prevalence could therefore be explained by the fact that wealthier individuals are more likely to travel to other cities with higher disease prevalence, such as Ouagadougou. Nevertheless, these results should not overshadow the fact that difficulties in accessing clean water have direct health repercussions beyond COVID-19 pandemic, particularly concerning protection against viruses and waterborne diseases. Unfortunately, in some developing countries, water scarcity has been exacerbated due to successive lockdowns, closures, or limited access to public water sources such as fountains and reservoirs. In our sample, only 7% of households had access to piped water at home or from neighbors. Note that the odd ratios of the other risk factors remain unchanged if access to water is excluded from the model (see Appendix Table S1).

Study Strengths: The study benefited from a large sample size and a low refusal rate. The population-based sample included individuals without targeting symptomatic or diagnosed at-risk populations.

Study Limitations: The main limitations of the study were the period of data collection (too late after the second wave to detect people infected during that peak) and the diagnostic performance of the rapid test used in an African population. One concern of the study is the lack of a clear understanding of the Biosynex COVID-19 BSS and other rapid tests’ diagnostic performance. The Biosynex COVID-19 BSS has been shown to have 91.8% (100%) sensitivity and 99.2% (99.5%) specificity for the IgM ( IgG) by the reference center of Institut Pasteur when using RT-PCR tests as the reference method for clinical diagnosis in 446 blood samples for the IgG and 456 blood samples for the IgM (the report is available upon request). However, these samples were collected from French patients and diagnostic performances might differ in African populations, in particular due to population-specific cross-reactivity. A study by Ouedraogo et al. estimated the specificity of the Biosynex COVID-19 BSS rapid test at 99.36% and sensitivity at 48.41%, compared to the WANTEI SARS CoV-2 Ab ELISA immunoassay as the reference test, suggesting an underestimation of seroprevalence in our study population [19]. Yet, it has also been shown that ELISA immunoassays can have relatively low specificity in West African populations [20, 21]. This raises concerns that estimating the sensitivity of rapid antibody tests from comparisons with ELISA immunoassays might underestimate the sensitivity of these tests. The extent of the underestimation of seroprevalence rates in our study therefore remains unclear in the absence of a validation of rapid antibody tests with RT-PCR as the reference method in our study population.

## Conclusions

The results of this study indicate that COVID-19 has been circulating more extensively within middle-sized cities in Burkina Faso than officially reported, given the low use of systematic screening. Prevention measures implemented to contain the pandemic must take these areas into consideration. Our findings highlight the vulnerability of women, older adults, and overweight and obese individuals to the epidemic.[4]

## Data Availability

All data produced in the present study are available upon reasonable request to the authors.

## Acknowledgments

The project received approval from the Burkina Faso Health Research Ethics Committee through deliberation n°2020-7-132.

We extend our gratitude to all participants who contributed to this study, dedicating their time to answer our questions, provide capillary blood samples for serological testing, and adhere to anthropometric measurements.

We address our acknowledgments to Macoura Doumbia, Muriel De Souza and Yara Mimboure for their contribution to data collection.

## Author Contributions

**Conceptualization:** Hermann Badolo, Abdramane Berthé, Philippe De Vreyer, Elodie Djemaï, Dramane Kania, Adama Sana, Thomas Thivillon.

**Data curation:** Philippe De Vreyer, Elodie Djemaï, Thomas Thivillon.

**Data analysis:** Elodie Djemaï, Adama Sana.

**Funding acquisition:** Hermann Badolo, Abdramane Berthé, Philippe De Vreyer, Elodie Djemaï, Dramane Kania, Adama Sana, Thomas Thivillon.

**Investigation:** Hermann Badolo, Abdramane Berthé, Philippe De Vreyer, Elodie Djemaï, Dramane Kania, Adama Sana, Thomas Thivillon.

**Methodology:** Hermann Badolo, Abdramane Berthé, Philippe De Vreyer, Elodie Djemaï, Dramane Kania, Adama Sana, Thomas Thivillon.

**Project administration:** Hermann Badolo, Abdramane Berthé, Philippe De Vreyer, Elodie Djemaï, Dramane Kania, Adama Sana, Thomas Thivillon.

**Software:** Philippe De Vreyer, Elodie Djemaï, Thomas Thivillon

**Supervision:** Philippe De Vreyer, Adama Sana.

**Writing – original draft:** Elodie Djemaï, Adama Sana.

**Writing – review & editing:** Philippe De Vreyer, Elodie Djemaï, Adama Sana, Thomas Thivillon. All authors agree that the paper is submitted to PLoS ONE.

## Supplementary material

S1 File. This is the S1 File Title: Table S1: Odds ratio from multivariate logistic regression (not controlling for access to water).

## Notes

### Competing Interest Statement

The authors have declared no competing interest.

### Funding Statement

This study was funded by ANRS MIE, an antonomous agency of Inserm, and by the French Development Agency (AFD).

### Author Declarations

The Health Ethics Research Committee of the Ministry of Health of Burkina Faso gave approval for this work.

