## Appendix Table S1 for "Seroprevalence and risk factors for SARS-CoV-2 infection in middle-sized cities of Burkina Faso: a descriptive cross-sectional study"

**Table S1:** **Odds ratio from multivariate logistic regression (not controlling for access to water)**

| **Variables** | **Multivariate OR** | **95pc-CI** | **P-value** |
|---|---|---|---|
| **Female** | 1.47 | 1.11-1.96 | 0.008 |
| **BMI equal to 25 and above** | 1.46 | 1.07-1.98 | 0.015 |
| **55 years old or older** | 1.60 | 1.17-2.18 | 0.003 |
| **Constant term (baseline odds)** | 0.08 | 0.07-0.11 | 0.000 |
| **Number of observations** | 2,141 | | |

*Notes*. Sample : Household members aged 16 or older whose RDT is valid. BMI body mass index, n number of individuals, OR odds ratio, CI confidence interval.
